# Biomechanical Strain Responses in the Optic Nerve Head Region in Glaucoma Patients After Intraocular Pressure Lowering

**DOI:** 10.1101/2023.05.30.23290662

**Authors:** Zhuochen Yuan, Cameron A. Czerpak, Michael Saheb Kashaf, Harry A. Quigley, Thao D. Nguyen

**Affiliations:** Wilmer Ophthalmological Institute, Johns Hopkins University School of Medicine, Baltimore, MD 21287, USA.; Department of Mechanical Engineering, The Johns Hopkins University, Baltimore, MD, 21218, USA.

**Keywords:** glaucoma, optic nerve head, strain, intraocular pressure, sclera.

## Abstract

**Objective:** To measure strain values and their association with intraocular pressure (IOP) change across five posterior eye regions in glaucoma patients.

**Design:** Cohort study.

**Participants:** Glaucoma patients who were imaged with optical coherence tomography (OCT) prior to and after laser suturelysis following trabeculectomy surgery (29 image pairs, 26 persons)

**Intervention:** Noninvasive imaging of the eye.

**Main Outcomes:** Strain values in eye regions.

**Results:** Mean strains were lowest in the retina and highest in the prelaminar neural tissue (PLNT) for *E_max_*, *Γ_max_*, and *E_zz_*. The values of *E_max_* in the anterior lamina cribrosa (ALC) and sclera were significantly related (P=0.0094, linear regression). Higher axial strain (*E_zz_*) was significantly associated with greater IOP decrease in the ALC, PLNT, and retina (P<0.05). Higher *Γ_max_* and *E_max_* strains were significantly associated with greater IOP decreases across all 5 eye regions. ALC and PLNT had negative median radial (*E_rr_*) compliance, while sclera had positive *E_rr_* compliance (P=0.017). *E_max_* and *Γ_max_* strains of the ALC were significantly and positively associated with these strains in the other 4 regions (P<0.005). Likewise, the *E_zz_* of ALC had a significant positive relationship with the other 4 regions (P<0.05).

**Conclusions:** Regional strains in the optic nerve head zone can be effectively measured using OCT and are related to the magnitude of IOP change. Strains were largest in PLNT and ALC and were smallest in retina. The sclera and choroid on average expand radially and circumferentially indicating a volume increase with IOP lowering.

## Introduction

The mechanical behavior of tissues in the posterior eye, especially in the optic nerve head (ONH) region determines their response to intraocular pressure (IOP)^1^, which is a major factor in axon injury in glaucoma.^2^ Modeling studies indicate that the behavior of the ONH, especially the anterior lamina cribrosa (ALC) and peripapillary sclera (PPS), are vital to the susceptibility to glaucoma damage.^3–6^ Biomechanical strains of the ALC and sclera have been studied in *post mortem* human eyes,^7–11^ using inflation tests with laser scanning multiphoton imaging and digital volume correlation of the images to estimate the pressure-induced strain response.^12–16^

To estimate ALC strains in the living eye, Girard and co-workers,^17–19^ Fazio and co-workers,^20, 21^ Sigal and co-workers,^22, 23^ and Nguyen and co-workers^24, 25^ combined optical coherence tomography (OCT) imaging of the LC and digital volume correlation (DVC) image analysis to derive ALC strain measurements after short-term IOP change. When IOP was increased by ophthalmodynamometer pressure on the sclera for 2 minute periods, significant ALC strains were measured.^26, 27^ In some open angle glaucoma (OAG) patients,^18^ ALC strains with such short-term IOP increase were greater in eyes with worse visual field sensitivity. Using images taken 20 minutes after IOP-lowering by suturelysis after trabeculectomy,^25^ we found that ALC strains were on average tensile (expanding) in the anterior-posterior axis and compressive radially. Similar to Chuangsuwanich et al.^18^, we found that eyes with worse glaucoma damage had greater strain for each unit of IOP lowering.

This study builds upon these findings by analyzing the association between strains and IOP decrease in multiple regions of the posterior eye, including the prelaminar neural tissue (PLNT), retina, choroid, and sclera. The ultimate aim of these investigations is to develop greater understanding of the *in vivo* deformation response of the ONH tissues to IOP change and to develop clinically useful biomarkers for susceptibility to glaucoma damage.

## Methods

### Experimental subjects

The research was approved and supervised by the Institutional Review Board of the Johns Hopkins School of Medicine and abided by the tenets of the Declaration of Helsinki and can be found at ClinicalTrials.gov Identifier: NCT03267849. Written informed consent was obtained for each subject. Primary open angle glaucoma subjects were tested as part of a regularly scheduled visit to the Wilmer Institute Glaucoma Center of Excellence. Some data from the ALC of these patients has been previously published.^25^ They underwent OCT imaging immediately before and a mean of 20 minutes after suturelysis that was performed at post-operative visits following trabeculectomy surgery. One patient had suturelysis in both left and right eyes and two patients had repeated suturelysis in the same eye. This yielded 29 image pairs of 27 eyes from 26 patients.

The IOP was measured with an iCare tonometer (iCare Finland Oy, Espoo, Finland) immediately prior to each imaging session and the recorded IOP was the average of two mean measurements of 6 IOPs each. Previously published data on these patients show that the applanation tonometer values at baseline were not significantly different from the baseline iCare values. The data describing the patients is in Table 1.

**Table 1:**
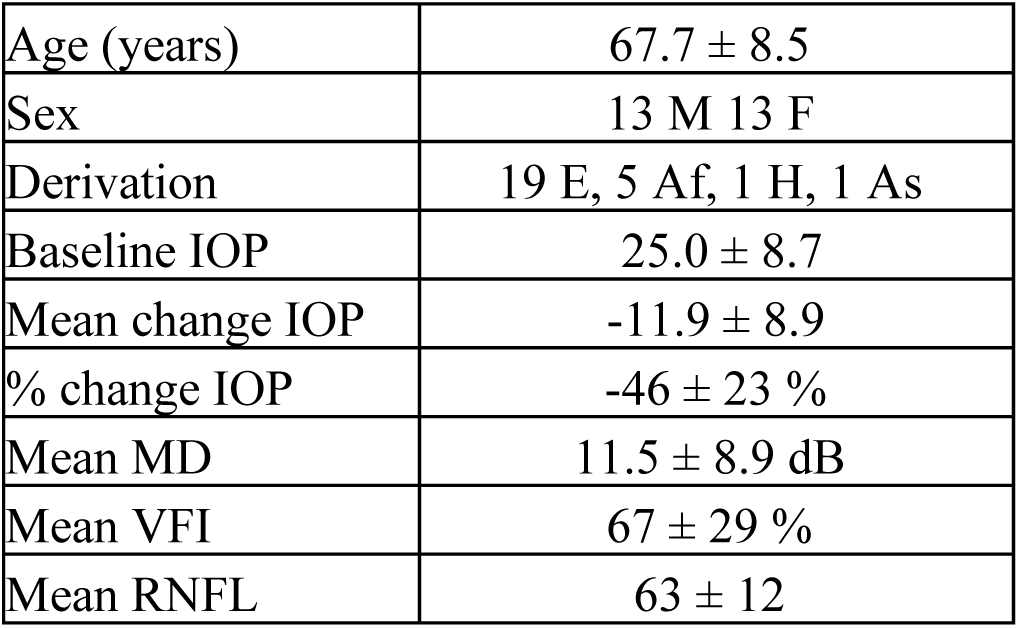
Patient Characteristics.

### OCT Imaging

Imaging was performed with the Heidelberg Spectralis (Heidelberg Engineering GmbH, Heidelberg 69121 Germany). Keratometry readings and axial length were measured using the IOL Master instrument (Zeiss, Dublin, CA) and keratometry measurements were entered prior to each imaging set. First, images were collected prior to suturelysis. Approximately 20 minutes after the suturelysis was performed, IOP and keratometry were remeasured and the second set of images was taken. For each session, 24 radial OCT scans were acquired in enhanced depth imaging and analysis carried out as published in detail.^25^

A masked observer (M.S.K.) manually marked the two ends of Bruch’s membrane on each side of the optic nerve head (ONH), the retinal pigment epithelium, the posterior border of the choroid, and the anterior border of the ALC at the baseline IOP (Figure 1).^28^ Manually marked ALC border positions were fit by piecewise linear interpolation to generate a smooth ALC boundary. Clear visibility of the BMO is crucial to this process, hence our choice of utilizing radial scans which have better BMO visibility in comparison to raster scans. To segment the different tissue regions, the lateral borders of the ALC were defined by two vertical lines projected from the ends of Bruch’s membrane. The posterior border of the ALC was defined by a curvilinear line parallel to the marked anterior ALC border and 250 µm posterior to it (Figure 1). The posterior lamina cribrosa (PLC) was defined as the zone 250 µm posterior to the ALC with a posterior limit parallel to the ALC boundary. The prelaminar neural tissue (PLNT) zone was defined as the tissue anterior to the ALC and internal to the vertical line drawn from the ends of Bruch’s membrane. The retina was defined as the tissue anterior to the pigment epithelium. The choroid was taken from the pigment epithelium to the marked choroidal—scleral interface. The sclera was all image data from below the choroidal scleral interface to the line vertically drawn down from the ends of Bruch’s membrane. However, strains were only obtained in regions with high signal to noise ratio, meaning that scleral strains were on average within 150µm of the choroidal scleral interface (Figure 1 B left sclera). Ideal signal to noise ratio allowed for strains to be identified deeper into the sclera (Figure 1 B right sclera).

**Figure 1:**
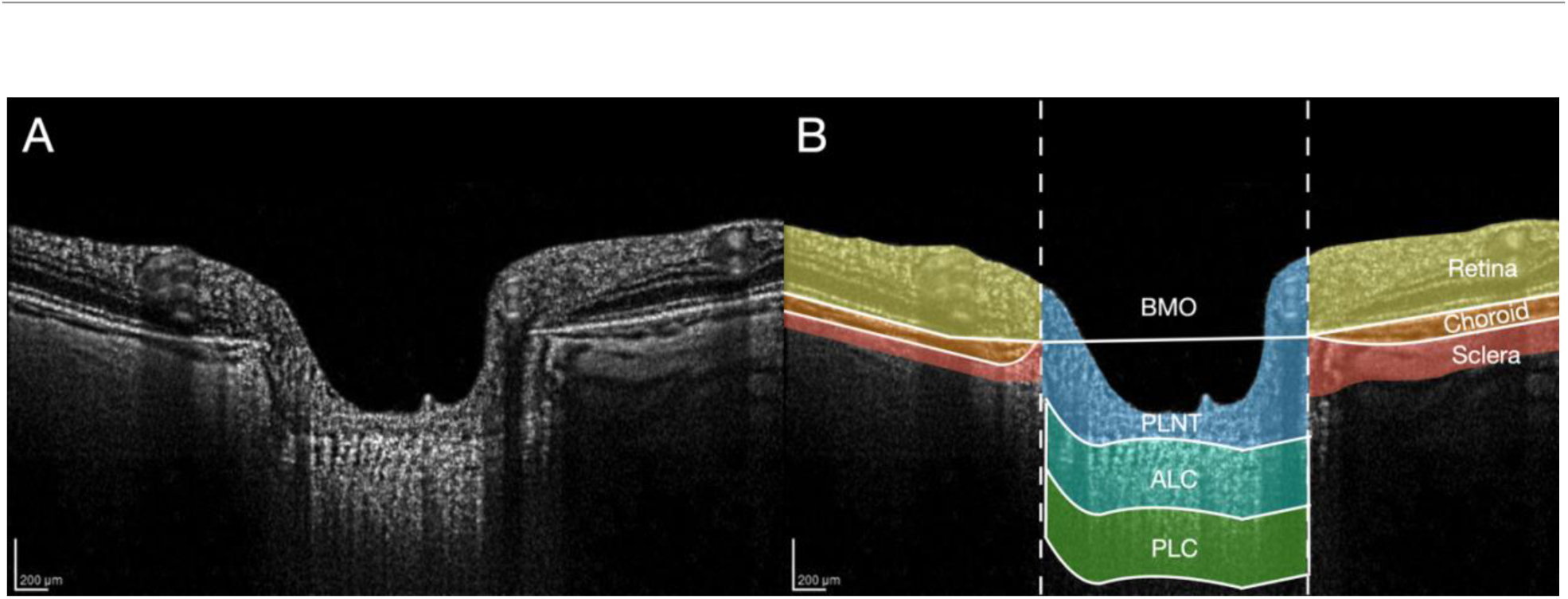
A, Slice 1 of the 24 radial OCT images following enhancement. B, Segmentation of the 6 tissues. The ALC and PLC were defined by regions 250µm in depth. Strains were only identified in regions with accurate DVC correlation. The left sclera (B left) was representative area of modest DVC correlation, while the right sclera (B right) had a larger area of accurate DVC correlation. ALC = anterior lamina cribrosa, PLC = posterior lamina cribrosa, BMO = Bruch’s membrane opening, PLNT = prelaminar neural tissue, DVC = digital volume correlation.

For retina, choroid and sclera, the image data from both sides of the optic nerve head were combined. Thus, the area to be analyzed varied by region. Areas reported are the total area summed across all 24 radial scans. For tissues with left and right sides (retina, choroid, and sclera), areas represent both sides added together. The anterior lamina depth (ALD) was defined as the mean distance from a line connecting the ends of Bruch’s membrane to the ALC anterior border, with change in ALD being calculated by the software utilized.

The details for the application of digital volume correlation (DVC) methodology to derive strains have been published in detail.^24, 25^ The method calculates the displacements and strains by correlating the unique patterns of image intensity between the two imaging volumes at different IOP. The areas of accurate DVC correlation are defined as those where the correlation coefficient was greater than a threshold of 0.055 and the displacement errors were less than 5 µm (0.7-1.3 pixels). Displacement outliers were also removed if they were 10 µm larger than the average of the displacements in an 8-pixel radius within the imaging plane. The image contrast in areas with a correlation coefficient below the threshold was too low to discern unique image patterns for displacement tracking. To calculate the baseline displacement errors, we applied DVC to correlate two images taken back-to-back before IOP lowering by suturelysis. In addition, we calculated the DVC correlation error by applying numerically a deformation to one of the two images taken before surgery, and applying DVC correlation to the deformed and undeformed image pairs. We calculated the area of accurate DVC correlation for the ALC, PLC, PLNT, retina, choroid, and sclera.

### Assessment of OCT and visual fields

OCT retinal nerve fiber layer (RNFL) imaging was used to define structural loss in glaucoma subjects, with at least 7/10 quality rating (Zeiss Cirrus OCT, Zeiss Meditec, Dublin, CA). Visual fields were performed on Zeiss 24-2 Sita Standard field tests (Zeiss HFA 2i, Zeiss Meditec, Dublin, CA). Past progressive change in OCT or field tests from the years prior to imaging was assessed by the Structure/Function GPA analysis of Zeiss Forum software. To qualify as prior change, the OCT or field progression designation of “possible” or “likely” had to be present on at least two tests without reverting back to no progression at the most recent pair of tests.

### Outcomes

DVC used OCT images taken before suturelysis as the reference state. We calculated averaged ALD, and strain components in a cylindrical coordinate system, *E_zz_*, *E_θθ_*, *E_rr_*, *E_rθ_*, *E_θz_*, *E_rz_*. We also calculated the maximum principal strain *E_max_* and the maximum shear strain *Γ_max_*, which signify the maximum tensile strain and maximum shear strain, respectively, over all orientations in the R-Z plane (plane of the radial scan). While all other strains were calculated in the 3d, E_max_ and Γ_max_ were calculated in the 2d plane of the radial scan due to the lower resolution of the out-of-plain strains *E_θθ_*, *E_rθ_*, and *E_zθ_*. The components *E_zz_*, *E_rr_*, *E_rz_* are the anterior-posterior strain, radial strain, and the in-plane shear strain, respectively, while *E_θθ_*, *E_rθ_*, and *E_θz_* are the circumferential (hoop) strain, twist strain, and out-of-plane shear. A normal strain deforms a square into a rectangle. Positive normal strains represent expansion in the stated direction (tensile), while negative strains represent contraction (contractile). Shear strains *E_rθ_* (radial-circumferential strain), *E_θz_* (circumferential-anterior-posterior strain), *E_rz_* (radial-anterior-posterior strain) represent shape distortion. A shear strain deforms a square into a rhombus. ALD is defined as the displacement of the anterior LC surface relative to the displacement of a line connecting the 2 points marking Bruch’s membrane opening. Positive ALD is a more posterior displacement of the ALC surface relative to Bruch’s membrane. We calculated the compliance for each outcome by dividing by IOP change (either change in mmHg or percent change from baseline). Data from all 29 test pairs were utilized, along with the effects on strain of visual field status, and field progressive change in the period prior to testing. RNFL measurements were obtained for each eye undergoing suturelysis. In one of the 29 eyes, a non-concurrent OCT RNFL thickness measurement was used to estimate thickness, which was at the typical floor value indicating severe damage.

### Statistical Analysis

The different strain values, *E_zz_*, and *E_ma_*_x_ and *Γ_max_* were compared to IOP change, while their compliance values were compared to average RNFL thickness and visual field indexes across the regions of the eye. These relationships were examined using a linear regression analysis through RStudio. Multivariate linear regression analysis used Matlab 2021a.

Compliance of the strain response was defined by dividing the strain value by the absolute value of IOP change or percent change in that eye. Statistical significance was indicated by p values < 0.05.

## Results

### 1. Correlation areas by region

The mean area available for analysis for each region was greatest in retina and PLNT and lowest in PLC (Table 2). The mean percent DVC correlation in the ALC region was 59 ± 16%, while in the PLC was only 10 ± 8%. Given the low value for available area for analysis in the PLC and its low percent correlation, no further data were presented for this zone. The height of the PLNT, retina, choroid, and sclera varied from eye to eye. Therefore, we could not calculate percent DVC correlation using the same method as the ALC and the PLC. We instead estimated the percent area correlation by measuring the average height and width of the retina and choroid of 3 representative eyes to estimate the total potential area that could have accurate DVC correlation. The area of accurate DVC correlation in the sclera for most eyes was a 100 - 200 μm thick region below the choroidal-scleral interface (Fig. 1). For the sclera, we measured the average width of 3 representative eyes, but assumed the height of the sclera was the top end of our range (200 μm) for the total potential area that could have accurate DVC correlation. Due to the irregular shape of the PLNT, we segmented 12 representative eyes to calculate the total area where accurate DVC correlation was possible. These areas were used to estimate the percent area correlation in the PLNT, retina, choroid, and sclera (Table 2).

**Table 2:**
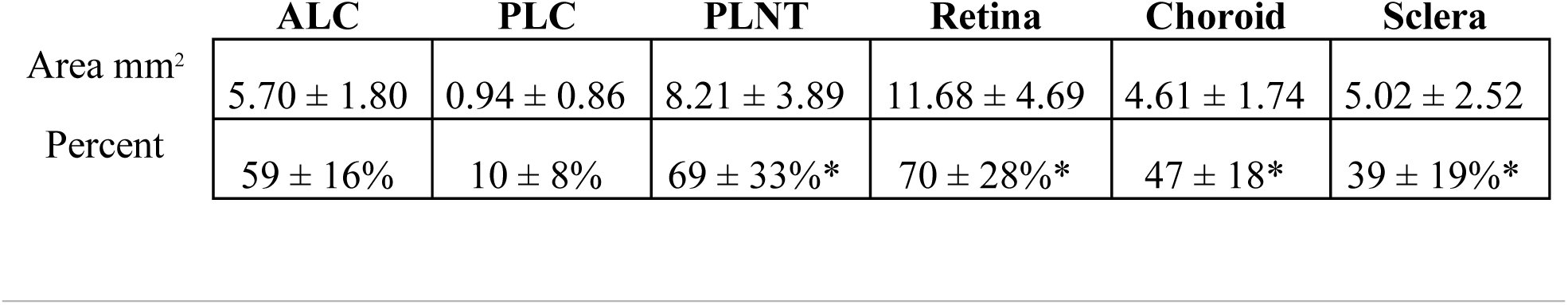
Mean area and percent correlation analyzed by region * = values estimated from representative eyes.

### 2. Baseline and correlation error estimates

The reproducibility (baseline error) of two sets of images taken immediately after each other and the mean correlation errors are presented in Table 3. The values for each region for both error types were tested to determine if the values were significantly different from zero error. None of the baseline errors were greater than no difference, while the correlation errors for *E_rr_*, *E_zz_* and *E_θθ_* were small, but greater than zero.

**Table 3:** Optic Nerve Head Baseline and Correlation Errors.

| Mean Baseline Errors |  |  |  |  |  |  |
| --- | --- | --- | --- | --- | --- | --- |
| | $E_{rr}$ | $E_{zz}$ | $E_{rz}$ | $E_{\theta\theta}$ | $E_{r\theta}$ | $E_{z\theta}$ |
| mean | 0.045% | 0.129% | 0.053% | -0.015% | 0.052% | -0.054% |
| stdev | 0.270% | 0.360% | 0.159% | 0.215% | 0.476% | 0.423% |
| median | -0.002% | 0.060% | 0.035% | 0.010% | -0.030% | 0.008% |
| p value | 0.37 | 0.64 | 0.08 | 0.71 | 0.56 | 0.50 |
| Mean Correlation Errors |  |  |  |  |  |  |
| | $E_{rr}$ | $E_{zz}$ | $E_{rz}$ | $E_{\theta\theta}$ | $E_{r\theta}$ | $E_{z\theta}$ |
| mean | -0.074% | 0.468% | 0.002% | 0.299% | 0.005% | 0.020% |
| stdev | 0.060% | 0.134% | 0.025% | 0.041% | 0.039% | 0.095% |
| median | -0.071% | 0.470% | -0.001% | 0.286% | -0.003% | 0.010% |
| p value | <0.0001 | <0.0001 | 0.59 | <0.0001 | 0.53 | 0.27 |

### 3. Mean Strains

Mean strains were lowest in the retina and highest in the PLNT for *E_max_*, *Γ_max_*, and *E_zz_* (Table 4). The *Γ_max_* strains were somewhat lower than the *E_max_* strains for each region. The mean *E_zz_* value was positive, indicating tensile strain (i.e., thickening) for all regions with IOP lowering. *E_θθ_* and shear strains did not achieve a mean value greater than correlation error in some regions (shaded values in Table 4).

**Table 4:** Mean strains by region. Shaded boxes indicate mean values below the mean correlation error for that strain/region.

| Regional Strains: Mean ± Standard Deviation | | | | | |
| --- | --- | --- | --- | --- | --- |
|  | ALC | PLNT | Retina | Choroid | Sclera |
| $E_{max}$ | 1.83 ± 1.54% | 1.97 ± 1.87% | 0.91 ± 0.62% | 1.21 ± 1.10% | 1.30 ± 0.96% |
| $\Gamma_{max}$ | 1.46 ± 1.14% | 1.46 ± 1.20% | 0.69 ± 0.36% | 0.81 ± 0.57% | 0.92 ± 0.54% |
| $E_{zz}$ | 0.94 ± 1.19% | 1.10 ± 1.79% | 0.35 ± 0.49% | 0.74 ± 1.03% | 0.57 ± 0.90% |
| $E_{rr}$ | -0.19 ± 0.33% | -0.09 ± 0.36% | 0.08 ± 0.22% | 0.06 ± 0.23% | 0.17 ± 0.33% |
| $E_{\theta\theta}$ | -0.21 ± 0.98% | -0.32 ± 0.57% | 0.001 ± 0.09% | 0.03 ± 0.07% | 0.04 ± 0.13% |
| $E_{r\theta}$ | 0.19 ± 0.01% | 0.10 ± 0.04% | 0.01 ± 0.001% | 0.004 ± 0.001% | -0.0004 ± 0.002% |
| $E_{rz}$ | 0.0003 ± 0.005% | 0.0003 ± 0.004% | -0.0005 ± 0.003% | -0.0005 ± 0.002% | 0.0003 ± 0.003% |
| $E_{z\theta}$ | -0.002 ± 0.006% | -0.002 ± 0.006% | 0.0001 ± 0.011% | 0.002 ± 0.01% | 0.003 ± 0.014% |

The values of *E_max_* in the ALC and sclera were significantly related to each other by linear regression (Figure 2, p = 0.0094, linear regression), as were the *E_max_* strains in ALC and PLNT (p<0.01), ALC and choroid (p = 0.006), and ALC and retina (p=0.001). Likewise, *Γ_max_* in ALC and sclera were linearly related (regression: y = 0.0041 + 1.13x, R^2^ = 0.29, p = 0.003), as was this strain in ALC and PLNT (p<0.001), ALC and choroid (p=0.002), and ALC and retina (p <0.001). *E_zz_* in ALC and sclera were not significantly associated (p = 0.071, linear regression), nor were *E_zz_* strains in the other 3 regions significantly related to that of ALC (all p>0.078).

**Figure 2:**
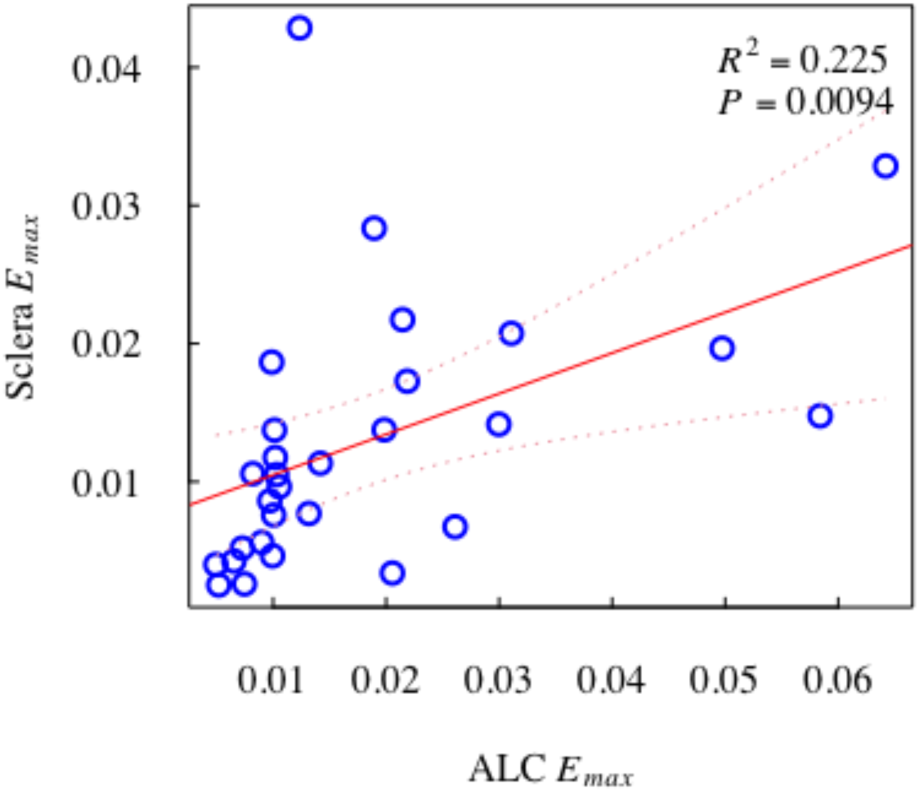
Relation of *E_max_* in ALC and Sclera

The *E_rr_* strain in the ALC and sclera were significantly related (p = 0.049, linear regression), but unlike the relationship in *E_max_*, for *E_rr_* the larger the tensile strain in sclera, the more compressive the strain in the ALC (Figure 3). None of the other strains that were measured had a significant relationship between the scleral strain and that of the ALC (all >0.05).

**Figure 3:**
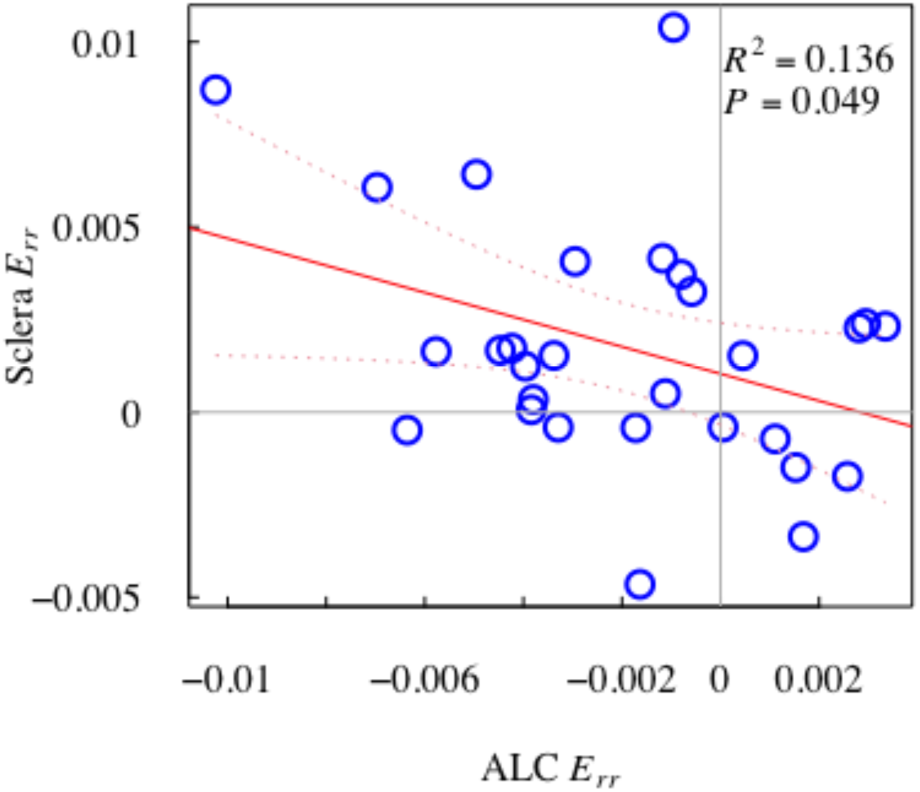
Relation of *E_rr_* Strain in ALC vs Sclera

### 4. Relationship of regional strains to IOP change

The *E_max_* and *Γ_max_* strains were significantly associated with both absolute IOP lowering and percent IOP lowering in each region, with greater percent IOP change leading to greater strain (p (retina) < 0.05, p (other regions) < 0.005, linear regression). The graphical relationships of *E_max_* in the ALC and choroid with percent IOP change are shown in Figure 4.

**Figure 4:**
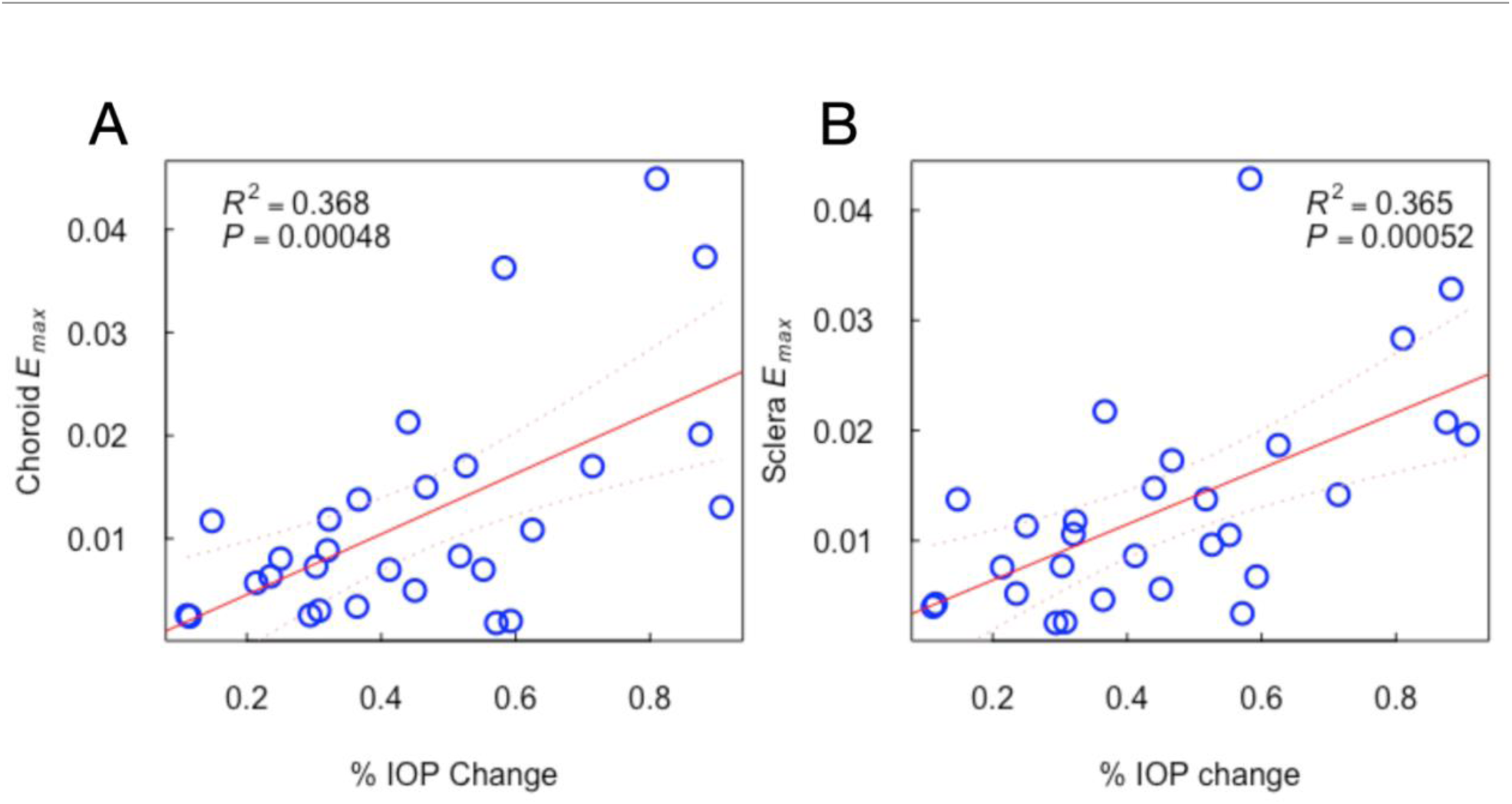
*E_max_* vs % IOP change in Choroid and Sclera

*E_zz_* was significantly greater with greater IOP change in the ALC (p = 0.018, linear regression), retina (p = 0.009), choroid (p = 0.0009), and PLNT (p = 0.007), but the relationship of scleral *E_zz_* to percent IOP lowering did not achieve significance (p = 0.13). Scleral E*_rr_* was significantly associated with percent IOP lowering (p = 0.0079), with greater E*_rr_* strains associated with greater percent IOP lowering as shown in Figure 5. There were no other strain components (i.e., *E_rr_*, *E_θθ_*) that had significant relationships to percent IOP lowering in any region (all >0.05)

**Figure 5:**
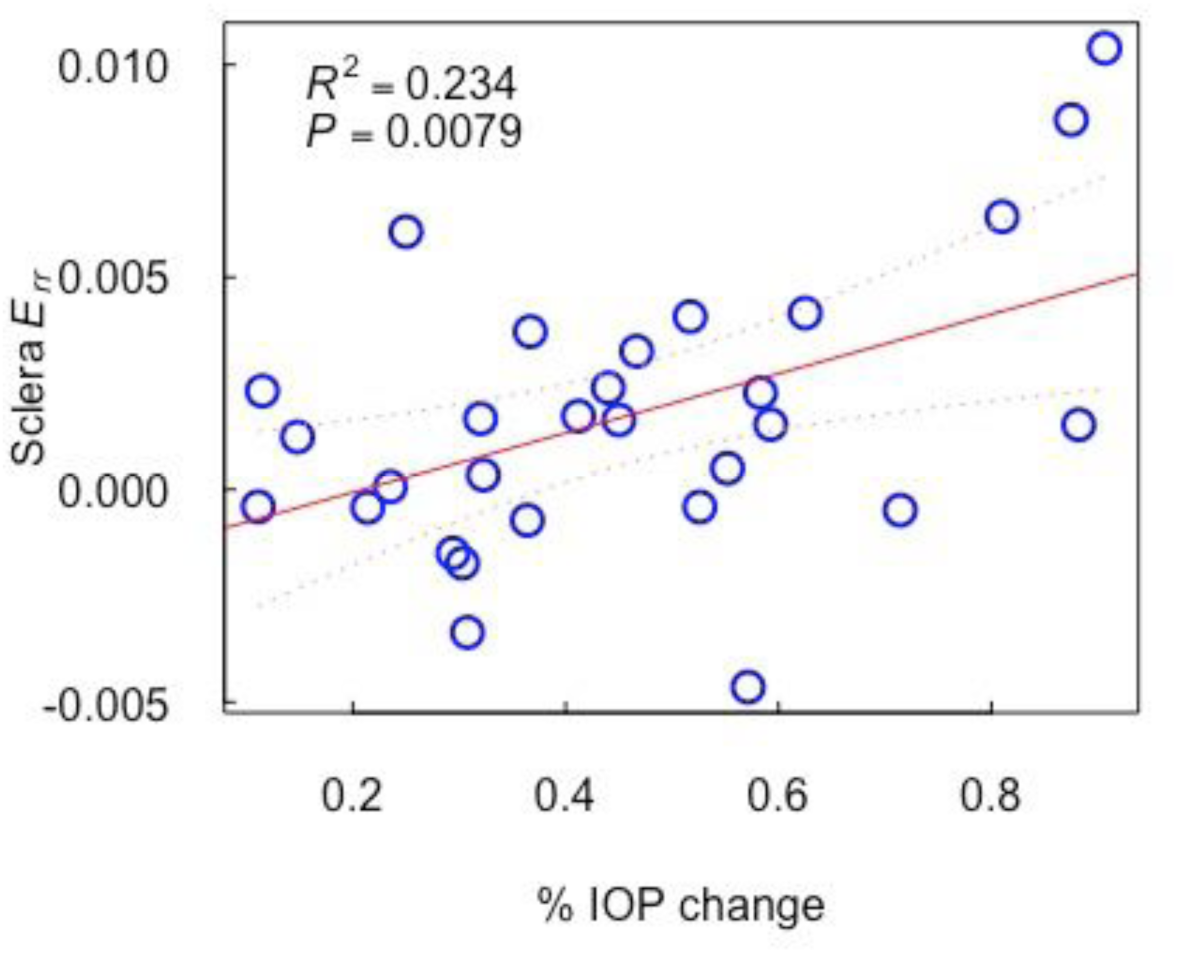
Sclera E*_rr_* vs % IOP change

### 5. Compliance by regions

We calculated the compliance for major strains as the ratio of the strain to the absolute and the percent IOP lowering (Table 5). The compliance calculated here is a structural property, which depends on the geometry and material properties. Compliance was highest in PLNT and ALC and lowest in retina and choroid, with the sclera intermediate in compliance. While the *E_zz_* compliance followed this order and were smaller in magnitude, all *E_zz_* compliance had median values significantly greater than zero, except for the retina (all but retina p<0.05, single sample t tests, Bonferroni corrected). The compliance values for *E_rr_* did not follow this same order, with the ALC and PLNT having negative median *E_rr_* compliance and the sclera having the largest positive compliance. Among *E_rr_* strains, only the scleral compliance was significantly greater than zero (p = 0.017). Compliance in other strains was not greater than zero.

**Table 5:** Median compliance (%strain/%IOP lowering) by tissue region

|  | **<u>Regional Compliance: Median Values</u>** |  |  |  |  |
| --- | --- | --- | --- | --- | --- |
| (%strain/%IOP lowering) | **<u>ALC</u>** | **<u>PLNT</u>** | **<u>Retina</u>** | **<u>Choroid</u>** | **<u>Sclera</u>** |
| **<u>$E_{max}$</u>** (%/%) | 3.60 | 3.76 | 2.05 | 2.38 | 2.66 |
| **<u>$\Gamma_{max}$</u>** (%/%) | 2.90 | 2.92 | 1.55 | 1.72 | 1.98 |
| **<u>$E_{zz}$</u>** (%/%) | 1.67 | 2.30 | 0.37 | 0.85 | 0.75 |
| **<u>$E_{rr}$</u>** (%/%) | -0.29 | -0.12 | 0.28 | 0.12 | 0.36 |

When regional compliances were compared for each of 4 strain components (*E_max_*, *Γ_max_*, *E_zz_*, and *E_rr_*: Table 6), patterns of difference are observed. For these comparisons, a paired Wilcoxon test was applied to regions two at a time, with significance levels adjusted for 10 pairs of comparisons, leading to a corrected significance level of 0.005 (bold values in Table 6). *E_max_* compliance was significantly higher in ALC and PLNT than in retina and choroid, with their values exceeding that of the sclera only in uncorrected p value (italics in Table 6). Scleral compliance was higher than that of retina in *E_max_*. For *Γ_max_* compliance, ALC and PLNT were significantly higher than retina, choroid and sclera, while the sclera was higher than retina and choroid. *E_zz_* compliance was higher in ALC, PLNT, and choroid than in retina. Scleral *E_zz_* compliance was borderline lower than ALC, PLNT and choroid, but was not significantly lower by corrected p values. *E_rr_* compliance was overall lower than *E_zz_*, and the most significant (uncorrected) difference was a larger compliance in the sclera than in the ALC, with the value in the latter being negative (Tables 5,6). The sclera was the only region in which the *E_rr_* compliance was significantly greater than zero (p = 0.017, t test), and only in this parameter was the sclera the highest region. There were no corrected significant differences between ALC and PLNT in any of the 4 compliance values, though *E_rr_* was borderline lower in PLNT than ALC (p = 0.075).

**Table 6:** Regional compliances comparisons using Wilcoxon test for each of 4 strain components (*E_max_*, *Γ_max_*, *E_zz_*, and *E_rr_*). Significance levels were adjusted for 10 pairs of comparisons such that p < 0.005 was significant (bold).

| **<u>Compliance Comparisons: p values</u>** |  |  |  |  |  |
| --- | --- | --- | --- | --- | --- |
| **<u>$E_{max}$</u>** | **<u>ALC</u>** | **<u>PLNT</u>** | **<u>Retina</u>** | **<u>Choroid</u>** | **<u>Sclera</u>** |
| **<u>ALC</u>** | - | <u>0.18</u> | **<u><0.0001</u>** | **<u>0.0017</u>** | <u>0.0075</u> |
| **<u>PLNT</u>** | - | - | **<u><0.0001</u>** | **<u>0.0038</u>** | <u>0.0060</u> |
| **<u>Retina</u>** | - | - | - | <u>0.024</u> | **<u>0.0002</u>** |
| **<u>Choroid</u>** | - | - | - | - | <u>0.039</u> |
| **<u>Sclera</u>** | - | - | - | - | - |
|  |  |  |  |  |  |
| **<u>$\Gamma_{max}$</u>** | **<u>ALC</u>** | **<u>PLNT</u>** | **<u>Retina</u>** | **<u>Choroid</u>** | **<u>Sclera</u>** |
| **<u>ALC</u>** | - | <u>0.94</u> | **<u><0.0001</u>** | **<u><0.0001</u>** | **<u>0.0013</u>** |
| **<u>PLNT</u>** | - | - | **<u><0.0001</u>** | **<u><0.0001</u>** | **<u>0.0003</u>** |
| **<u>Retina</u>** | - | - | - | <u>0.21</u> | **<u>0.0006</u>** |
| **<u>Choroid</u>** | - | - | - | - | **<u>0.0023</u>** |
| **<u>Sclera</u>** | - | - | - | - | - |
|  |  |  |  |  |  |
| **<u>$E_{zz}$</u>** | **<u>ALC</u>** | **<u>PLNT</u>** | **<u>Retina</u>** | **<u>Choroid</u>** | **<u>Sclera</u>** |
| **<u>ALC</u>** | - | <u>0.14</u> | **<u>0.0015</u>** | <u>0.12</u> | <u>0.038</u> |
| **<u>PLNT</u>** | - | - | **<u>0.0044</u>** | <u>0.11</u> | <u>0.076</u> |
| **<u>Retina</u>** | - | - | - | **<u>0.0005</u>** | <u>0.031</u> |
| **<u>Choroid</u>** | - | - | - | - | <u>0.79</u> |
| **<u>Sclera</u>** | - | - | - | - | - |
|  |  |  |  |  |  |
| **<u>$E_{rr}$</u>** | **<u>ALC</u>** | **<u>PLNT</u>** | **<u>Retina</u>** | **<u>Choroid</u>** | **<u>Sclera</u>** |
| **<u>ALC</u>** | - | <u>0.075</u> | <u>0.031</u> | <u>0.066</u> | <u>0.0069</u> |
| **<u>PLNT</u>** | - | - | <u>0.25</u> | <u>0.38</u> | <u>0.066</u> |
| **<u>Retina</u>** | - | - | - | <u>0.23</u> | <u>0.072</u> |
| **<u>Choroid</u>** | - | - | - | - | <u>0.046</u> |
| **<u>Sclera</u>** | - | - | - | - | - |

### 6. Relationship of strains to ALD change

As previously described in Czerpak et al.^25^, the ALD moved both into and out of the eye on IOP lowering, with a mean value of 1.3 ± 6.3 µm (median 2.1 µm, range -18.2 to 14.1 µm). No regional *E_max_* values were related to ALD change (all p > 0.3). *Γ_max_* strain was related to ALD change only in the ALC (p = 0.014). The *E_zz_* strain in the PLNT and sclera increased significantly at greater positive values of ALD change (p = 0.01, 0.046, respectively; Figure 6). This indicated that when the anterior ALD border moved out of the eye (defined as positive ALD change), the axial strain in both PLNT and sclera were more tensile, while in eyes with ALD movement into the eye, *E_zz_* strain was more compressive in PLNT and sclera. *E_zz_* strain was not related to ALD change in retina, choroid or ALC.

**Figure 6:**
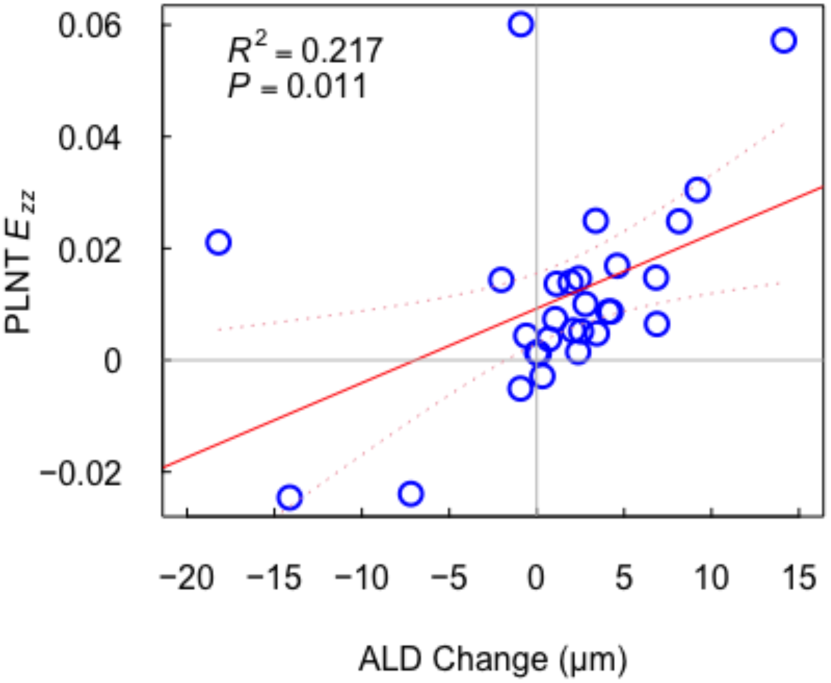
ALD change vs. *E_zz_* strain in PLNT

### 7. Compliances vs. RNFL, MD, and VFI for all regions

As previously reported for this cohort^25^, the *E_max_* and *Γ_max_* compliances for the ALC were significantly related to average RNFL thickness, with more abnormal RNFL associated with greater compliance (p = 0.049, 0.022). The *E_max_* and *Γ_max_* compliances for retina, choroid, PLNT, or sclera were not significantly related to either mean RNFL, MD, or VFI (all p>0.08, linear regression). No *E_zz_*, *E_rr_*, nor any other strain compliance values were significantly related to RNFL or visual field indexes (all p>0.15).

### 8. Relationship to prior visual field testing

Prior to imaging, patients had a median of 4 years of past visual field testing. Six eyes had no prior progressive worsening, 11 had probable or definite worsening, and 12 did not have sufficient numbers of past tests to determine progression. The mean ALC *E_max_* for non-progressors was 1.3 ± 0.7%, while for progressors it was 2.0 ± 1.6% (p = 0.33, t test). The mean sclera *E_max_* for non-progressors was 1.0 ± 0.54 and 1.7 ± 1.3% for progressors (p = 0.25, t test).

### 9. Relationship to axial length

Change in axial length was measured in 14 suturelysis procedures and was significantly related to IOP change or percent change (p < 0.004). Therefore, we only used IOP change or percent change for linear regression analysis. However, axial length measured prior to imaging was measured in 28 suturelysis procedures and was not related to IOP change or percent change (p > 0.4). We used multivariate linear regression to determine the effects of percent IOP decrease and axial length before suturelysis on the regional strain response (Supplement Tables S1-S5). Longer axial length before suturelysis was associated with more contractile *E_θθ_* (p=0.013) and more positive *E_rz_* (p=0.027) in the ALC. In the PLNT, longer axial length before suturelysis was associated with more contractile *E_θθ_* (p=0.026). This indicates that longer eyes experienced greater circumferential contraction per percent of IOP decrease in the ALC and the PLNT. The choroid experienced more expansive *E_rr_* for a longer axial length (p=0.053). Axial length was not associated with strains for the retina or the sclera.

## Discussion

Strains were measurable in each of the 5 ONH regions, though they were relatively small given the short 20 minutes time between the baseline image and the image at lower IOP. Indeed, some strain components in some regions were too small to be considered significant when compared to the estimated DVC correlation errors. Girard et al.^19^ applied similar methods to measure the strain response to IOP lowering weeks to months after trabeculectomy. They found that the mean minimum principal strain (maximum compressive strain) was 9.3% in the sclera, 8.45% in the PLNT and 6.75% in the LC. The maximum tensile strains were of similar magnitude. These are more than 4 times larger than measured here 20 minutes after IOP lowering by laser suturelysis. The strains measured in the study by Girard et al.^19^ likely included the long-term effects of viscoelasticity and remodeling of the ONH following IOP decrease.

While their study captures long term structural remodeling, our data focuses on short term effects which provide a more comprehensive understanding of optic nerve head remodeling over time. More recently, Chuangsuwanich et al.^18^ studied the ONH strains from IOP elevation produced by ophthalmodynamometry. The range of strain in the LC (2-12% effective strain) was also of similar magnitude to the strain in the sclera (2-10% effective strain).

The magnitudes of *E_zz_*, *E_rr_*, *E_max_*, and *ϕ′_max_* were largest in the PLNT and ALC and were smallest in the retina, with some retina strains failing to achieve mean values statistically different than zero. The strains components were significantly related to the magnitude of IOP lowering, supporting the validity of the strain estimates. In all regions except the sclera, the *E_zz_* strain was significantly more tensile with greater IOP lowering. Since the *E_zz_* in the sclera was statistically greater than baseline error, this apparent lack of significant association with IOP change does not appear to be because of DVC error. Rather the scleral response is likely to be more structurally complex than those of the LC, choroid, and retina. Keeping in mind the OCT’s were taken 20 minutes apart, this highlights how PLNT and ALC are more susceptible to short term IOP changes. Increased attention to these structures during diagnosis and treatment may be crucial in cases of acute IOP changes.

The tissues of the ONH and the peripapillary sclera are deformed by the intracranial pressure (ICP) in addition to the IOP. While IOP acts on the sclera and LC, the ICP only acts at the posterior surface of the LC and in the subarachnoid space. Computational studies and experimental studies have shown that an increase in IOP and ICP deform the peripapillary sclera and LC differently.^21, 29–33^ Moreover, the effect depends on the initial values of ICP and IOP in addition to the IOP and ICP change. The lack of association between the scleral strains and IOP may indicate a significant effect of ICP in this region. Alternatively, the range of IOP change in our data may not be sufficient to demonstrate changes in scleral strain in some strain components.

In this study, the mean *E_zz_* and *E_max_* were smaller in the sclera than in the ALC, but the difference was not statistically significant. The radial *E_rr_* and circumferential strains *E_88_* were also smaller in magnitude in the sclera than the ALC, but they were on average tensile rather than compressive as in the ALC. This means that the inner region of the sclera on average, thickens and expands radially and circumferentially with IOP lowering. This is in contrast to previous *ex vivo* inflation tests, which showed tensile mean *E_rr_* and *E_88_* strains in the sclera with IOP increase.^12, 34, 35^ We also performed *ex vivo* inflation tests of the human posterior scleral cup using laser scanning microscopy, second harmonic generation and two photon fluorescent imaging, and digital volume correlation to calculate the strains at higher resolution in the posterior regions of the LC and peripapillary sclera.^36^ While the mean *E_88_* in the peripapillary sclera was tensile in response to an IOP increase, the mean *E_rr_* in the peripapillary sclera was compressive for specimens with larger *E_rr_* in the LC. We speculated that greater bowing of the LC, as indicated by a larger tensile *E_rr_*, produced bending stress on the posterior surface of the peripapillary sclera at the boundary with the LC. The tensile *E_zz_*, *E_88_*, and *E_rr_* measured here for sclera indicates that the tissue volume increased with IOP lowering. The choroid also experienced a mean tensile *E_zz_*, *E_88_*, and *E_rr,_* indicating a volume increase with IOP lowering. The strain response of the inner region of sclera may be constrained by the compressible choroid. This, along with differences in the collagen structure between the inner and outer scleral regions,^37^ and differences in ICP would contribute to the differences in the *in vivo* strain measurements in the inner sclera and the *ex vivo* strain measurements in the outer sclera.

As previously noted, the ALD change can be either into or out of the eye with IOP lowering, but where regions had significant strains, they were tensile regardless of the direction of ALD movement. In the attempt to study LC tissue responses *in vivo*, many past reports used the position of the anterior LC border as a convenient surrogate for LC biomechanical responsiveness, because it is easily identified in spectral domain—optical coherence tomography (SD-OCT) imaging.^28, 38–43^ Our data again confirms that ALD as a dynamic measure is not, by itself, a good indicator of the state change in the ALC tissue. When the anterior ALD border moved out of the eye, the axial (*E_zz_*) strain in both PLNT and sclera were more tensile, while in eyes with ALD movement into the eye, *E_zz_* strain was more compressive in PLNT and sclera. *E_zz_* strain was not related to ALD change in retina, choroid or ALC. These data continue to point out that the position of the anterior border of the ALC is a poor parameter to judge tissue behavior in the ONH.

We previously showed that ALC *Γ_max_* strain was more compliant with greater loss of RNFL, but none of the other regions shared this relationship in that their compliances were not related to the degree of glaucoma injury estimated from RNFL or visual field index levels. A trend, but not a significant one, suggested eyes with past progression had greater strain. It will remain for more extensive testing, especially longitudinal follow-up of strain behavior, to determine the biomechanical parameters that are potential biomarkers for prospective glaucoma susceptibility. In that regard, we have developed methods to measure ONH region strains produced by addition or cessation of typical glaucoma eye drops that would provide the mechanism for strain measurements at baseline and over time in any glaucoma patient and in control subjects in research studies.

These data have some notable limitations. We recognize that OCT images fail to acquire usable data from a substantial minority of the ONH tissues. This derives in part from the blocking effect of overlying blood vessels and simply the attenuation of reflectance information by the thickness of the structures. Second, we made reasonable, but arbitrary choices about the information that would be defined as the ALC and the sclera. In the cases of both structures, the OCT images do not provide adequate contrast changes that demarcate their posterior limits. Our choice of the tissue included in the ALC is based on sound human histological measurements.

For the sclera, we are confident that the lack of penetration depth for the wavelength used encompasses only sclera, as it was, in general, only the first 100-200 μm of the tissue. However, there may be variations in scleral strain by depth that are not captured in our data. Our imaging method of choice is a radial scan which provides better definition of the Bruch’s membrane opening. However, radial scans encounter a unique problem as the distance between B scans is larger towards the periphery. This may lead to uneven 3d resolutions which can affect our analysis resulting in variations of strain in peripheral structures that are not fully captured by our data.

In conclusion, we present practical methods that can assess the biomechanical behavior of tissues in the ONH region. It will be important to measure strains in the PLNT, the ALC, and the sclera to elucidate important responses to IOP change. The movement of the anterior lamina cribrosa border is not related consistently to the strains within the lamina itself.

### Abbreviations

ALC: anterior lamina cribrosa
PLC: posterior lamina cribrosa
PLNT: pre-lamina neural tissue
ONH: optic nerve head
PPS: peripapillary sclera
OCT: optical coherence tomography
RNFL: retinal nerve fiber layer
MD: mean deviation
VFI: visual function index
IOP: intraocular pressure

## Data Availability

All data produced in the present study are available upon reasonable request to the authors

