## Supplemental Tables for "Biomechanical Strain Responses in the Optic Nerve Head Region in Glaucoma Patients After Intraocular Pressure Lowering"

Supplemental Material

### S1 Axial Length

Axial length measured prior to imaging was measured in 28 suturelysis procedures and was not related to IOP change or percent change (p > 0.04). Therefore, we used multivariate linear regression to determine the effects of percent IOP decrease and axial length before suturelysis on the regional strain response.

Supplemental Table S1: Multivariate linear regression analysis comparing percent IOP decrease and axial length before suturelysis to the strain in the ALC. Also shown are *p*-values and associated $R^2$ value (n = 28, 25 patients).

| | Value at zero IOP and axial length | *p*-value | Change per 1% IOP decrease | *p*-value | Change per 1μm axial length increase | *p*-value | $R^2$ |
|---|---|---|---|---|---|---|---|
| LC depth change (μm) | 8.41 | 0.736 | -0.0331 | 0.556 | -0.227 | 0.812 | 0.0148 |
| $E_{zz}$ | -0.0554 | 0.191 | 0.000257 | **0.0106** | 0.00209 | 0.198 | 0.2519 |
| $E_{rr}$ | -0.0115 | 0.368 | -3.80E-05 | 0.192 | 0.000444 | 0.367 | 0.111 |
| $E_{\theta\theta}$ | 0.0832 | 0.0212 | 6.71E-05 | 0.387 | -0.00347 | **0.0129** | 0.264 |
| $E_{r\theta}$ | 0.00317 | 0.874 | 8.05E-05 | 0.0841 | -0.000181 | 0.8143 | 0.124 |
| $E_{z\theta}$ | -0.0187 | 0.407 | -2.20E-05 | 0.663 | 0.000717 | 0.407 | 0.0405 |
| $E_{rz}$ | -0.0384 | 0.0326 | -7.33E-06 | 0.850 | 0.00154 | **0.0266** | 0.191 |
| $E_{max}$ | -0.0493 | 0.277 | 0.000479 | **<0.0001** | 0.00181 | 0.296 | 0.480 |
| $\Gamma_{max}$ | -0.0158 | 0.616 | 0.000370 | **<0.0001** | 0.000546 | 0.653 | 0.527 |

Supplemental Table S2: Multivariate linear regression analysis comparing percent IOP decrease and axial length before suturelysis to the strain in the PLNT. Also shown are *p*-values and associated $R^2$ value (n = 28, 25 patients).

| | Value at zero IOP and axial length | *p*-value | Change per 1% IOP decrease | *p*-value | Change per 1μm axial length increase | *p*-value | $R^2$ |
|---|---|---|---|---|---|---|---|
| LC depth change (μm) | 8.41 | 0.736 | -0.0331 | 0.556 | -0.227 | 0.812 | 0.0148 |
| $E_{zz}$ | 0.0226 | 0.720 | 0.000379 | **0.0125** | -0.00113 | 0.640 | 0.245 |
| $E_{rr}$ | 0.0101 | 0.478 | 4.34E-06 | 0.892 | -0.000444 | 0.419 | 0.0291 |
| $E_{\theta\theta}$ | 0.0457 | 0.0352 | -3.04E-05 | 0.516 | -0.00186 | **0.0260** | 0.185 |
| $E_{r\theta}$ | -0.0112 | 0.483 | 1.75E-05 | 0.624 | 0.000452 | 0.459 | 0.0276 |
| $E_{z\theta}$ | -0.0127 | 0.579 | -8.52E-05 | 0.105 | 0.000558 | 0.524 | 0.129 |
| $E_{rz}$ | -0.0210 | 0.1519 | 1.98E-05 | 0.541 | 0.000804 | 0.153 | 0.0849 |
| $E_{max}$ | 0.0221 | 0.674 | 0.000590 | **<0.0001** | -0.00114 | 0.572 | 0.523 |
| $\Gamma_{max}$ | 0.0057 | 0.859 | 0.000398 | **<0.0001** | -0.000352 | 0.775 | 0.564 |

Supplemental Table S3: Multivariate linear regression analysis comparing percent IOP decrease and axial length before suturelysis to the strain in the retina. Also shown are *p*-values and associated $R^2$ value (n = 28, 25 patients).

| | Value at zero IOP and axial length | *p*-value | Change per 1% IOP decrease | *p*-value | Change per 1μm axial length increase | *p*-value | $R^2$ |
|---|---|---|---|---|---|---|---|
| LC depth change (μm) | 8.41 | 0.736 | -0.0331 | 0.556 | -0.227 | 0.812 | 0.0148 |
| $E_{zz}$ | -0.00558 | 0.748 | 0.000108 | **0.00992** | 0.000167 | 0.802 | 0.239 |
| $E_{rr}$ | -0.0140 | 0.0751 | 3.55E-05 | **0.0469** | 0.000516 | 0.0868 | 0.206 |
| $E_{\theta\theta}$ | 0.00160 | 0.657 | -4.39E-06 | 0.589 | -5.32E-05 | 0.700 | 0.0155 |
| $E_{r\theta}$ | -0.00109 | 0.797 | 2.31E-05 | **0.0219** | -4.14E-06 | 0.980 | 0.198 |
| $E_{z\theta}$ | -0.0713 | 0.113 | -2.08E-05 | 0.833 | 0.00283 | 0.102 | 0.111 |
| $E_{rz}$ | 0.01019 | 0.361 | -2.21E-06 | 0.929 | -0.000418 | 0.329 | 0.0384 |
| $E_{max}$ | -0.02145 | 0.280 | 0.000174 | **0.000518** | 0.000889 | 0.244 | 0.393 |
| $\Gamma_{max}$ | -0.0117 | 0.306 | 0.000103 | **0.000397** | 0.000547 | 0.212 | 0.406 |

Supplemental Table S4: Multivariate linear regression analysis comparing percent IOP decrease and axial length before suturelysis to the strain in the choroid. Also shown are *p*-values and associated $R^2$ value (n = 28, 25 patients).

| | Value at zero IOP and axial length | *p*-value | Change per 1% IOP decrease | *p*-value | Change per 1μm axial length increase | *p*-value | $R^2$ |
|---|---|---|---|---|---|---|---|
| LC depth change (μm) | 8.41 | 0.736 | -0.0331 | 0.556 | -0.227 | 0.812 | 0.0148 |
| $E_{zz}$ | -0.0493 | 0.135 | 0.000289 | **0.000469** | 0.00172 | 0.173 | 0.401 |
| $E_{rr}$ | -0.0170 | 0.0500 | 2.50E-05 | 0.191 | 0.000645 | **0.0526** | 0.172 |
| $E_{\theta\theta}$ | 0.000874 | 0.756 | -1.47E-06 | 0.816 | -2.11E-05 | 0.845 | 0.00323 |
| $E_{r\theta}$ | -0.00209 | 0.563 | 2.07E-05 | **0.0165** | 4.78E-05 | 0.730 | 0.209 |
| $E_{z\theta}$ | -0.0438 | 0.251 | -2.31E-05 | 0.786 | 0.00182 | 0.214 | 0.0695 |
| $E_{rz}$ | 0.000963 | 0.918 | -1.11E-05 | 0.598 | -3.98E-05 | 0.912 | 0.0113 |
| $E_{max}$ | -0.0638 | 0.0611 | 0.000326 | **0.000154** | 0.00240 | 0.0656 | 0.460 |
| $\Gamma_{max}$ | -0.0306 | 0.0858 | 0.000169 | **0.000181** | 0.00122 | 0.0746 | 0.452 |

Supplemental Table S5: Multivariate linear regression analysis comparing percent IOP decrease and axial length before suturelysis to the strain in the sclera. Also shown are *p*-values and associated $R^2$ value (n = 28, 25 patients).

| | Value at zero IOP and axial length | *p*-value | Change per 1% IOP decrease | *p*-value | Change per 1μm axial length increase | *p*-value | $R^2$ |
|---|---|---|---|---|---|---|---|
| LC depth change (μm) | 8.41 | 0.736 | -0.0331 | 0.556 | -0.227 | 0.812 | 0.0148 |
| $E_{zz}$ | -0.0436 | 0.199 | 0.000124 | 0.107 | 0.00171 | 0.191 | 0.137 |
| $E_{rr}$ | -0.0107 | 0.3584 | 7.21E-05 | **0.00950** | 0.000355 | 0.424 | 0.243 |
| $E_{\theta\theta}$ | 0.00679 | 0.189 | -7.04E-06 | 0.540 | -0.000239 | 0.228 | 0.0639 |
| $E_{r\theta}$ | -0.00518 | 0.493 | 1.92E-05 | 0.263 | 0.000163 | 0.572 | 0.0553 |
| $E_{z\theta}$ | -0.0318 | 0.519 | -0.000140 | 0.215 | 0.00158 | 0.406 | 0.0994 |
| $E_{rz}$ | -0.0141 | 0.183 | 1.08E-05 | 0.647 | 0.000556 | 0.173 | 0.0748 |
| $E_{max}$ | -0.0537 | 0.0718 | 0.000273 | **0.000265** | 0.00212 | 0.0641 | 0.439 |
| $\Gamma_{max}$ | -0.0265 | 0.0888 | 0.000175 | **<0.0001** | 0.00109 | 0.0690 | 0.527 |
